# [Knowledge and attitudes regarding infant BLS among babysitters and nursemaids in north rural Jordan]

**DOI:** 10.1101/19014175

**Authors:** Liqaa Raffee, Shereen Hamadneh, Manal Kassab, Fidaa Almomani, Jehan Hamadneh, Mohammed AlBashtawy, Rawan A. Obeidat, Othman Almazloom

## Abstract

**Background:** There is a lack of national data related to the public knowledge and attitudes concerning Basic Life Support (BLS) for an infant. This study is the first to assess the knowledge and attitude regarding infant BLS among babysitters and nursemaids who are working in registered nurseries in North Jordan.

**Methods:** A semi-structured questionnaire was conducted among a convenience sample of 77 child caretakers (babysitters and nursemaids) who were looking after infants in registered nurseries in north of Jordan.

**Results:** The findings show that the majority of participants were not aware of skills for managing foreign body aspiration (or CPR) among infants, while more than two-thirds of them never attended training and or education sessions regarding managing foreign body aspiration among infants.

**Conclusions:** This research strongly recommends training for babysitters and care center staff. The findings of this study indicated that caregivers in Jordanian kindergartens and nurseries lack awareness regarding safe infant care. As such, there is a lack of basic skills for resuscitation in urgent cases; which can increase the risk of sudden and unexpected child death. Accordingly, the infant life support course is highly recommended for babysitters and nursemaids. Health care professionals could help to implement infant basic life support training courses through local community basic service centers, non-governmental organizations, local associations, universities and health colleges through providing workshops that aim at enhancing knowledge and skills for infant BLS among caregivers and babysitters.

## Introduction

Approximately 11% of children die each year as a result of a variety of causes including road traffic accidents (RTAs), falls, blast injuries, burns, insect bites, drowning, suffocations, and aspiration.(1) Pediatric cardiac arrest is a life-threatening event, which often is required a fast effective intervention.(2,3) There are more than 5,000 cases of pediatric cardiac-arrests out-hospital annually.(2,3) In particular, one-fifth of total child deaths were due to mechanical suffocation where the child was younger than four years.(3, 4) Dealing with infants’ urgent life-threatening situations and providing life support measures may improve infant survival rate. Moreover, the quality of the CPR administered is of paramount importance because it directly affects the infants’ hemodynamics, their survival, and the neurological outcomes after the arrest.(3)

The aspiration of the foreign body is a potentially life-threatening event especially in those under the age of 5 years.(5) An aspirated child can be presented to the emergency as a spectrum, ranging from an event that has no symptoms to more severe events including suffocation, cyanosis, respiratory distress, or even death.(5-7) However, in some cases, previous foreign objects that were topical in the lung can travel back to the mouth and are swallowed, after some severe cough episodes.(7)

The American Academy of Pediatrics has recommended that children’s caregivers should have life support skills and be trained to deal with life-threatening situations.(8) Survival after cardiopulmonary arrest is depending on the effectiveness of the intervention which includes the early management, CPR quality and the time to defibrillation.(9) Babysitters and the local community in Jordan lack awareness regarding safe infant care and basic skills for urgent cases which can increase the risk of sudden and unexpected child death.(10)

### Background

Choking and foreign body aspiration remain among the major causes of mortality and morbidity in young children, however, it could be preventable.(4, 5, 6, 11-14) Anyone can be a lifesaver for a cardiac arrest victim with the proper awareness and training.(10, 14, 15) Primary and secondary infant caregivers including parents, babysitters, and nursemaids should have the basic skills for life support to be able to use it when the baby’s life is at stake. (10-14) Young children aging between 0 and 3 years, are at the highest risk for accidental suffocation, due to several factors; young children tend to put small things in their mouths and often crying, screaming, running, and playing while such objects are still in their mouths; all of which could be accidentally ingested and cause suffocation. (4) In addition, they do not have molar teeth to chew some foods completely. (4) The most common reported cases of foreign body aspiration are organic or food related, especially when the child begins to eat solid food. (4)

CPR accurate application depends on experiences, skills, training, and confidence of the rescuer.(12) The American Heart Association released revised CPR recommendations in 2010 which state that the compression to breathing ratio is 30:2 for children with one rescuer, and 15:2 for children with two rescuers.(12)

Public awareness campaigns and the strict guidelines for toy manufacturers to be within safety requirements in the developed countries have contributed to decreasing the number of childhood deaths due to foreign body aspiration over the past decade.(11) However, in developing countries like Jordan, the public knowledge and skills related to infant basic life support and safety care are still not well established.(1, 10, 13)

### Study significance

In Jordan, there is a lack of data related to the public knowledge and attitudes towards Basic Life Support (BLS) for an infant. Therefore, assessing the knowledge and attitudes regarding infant BLS among babysitters and nursemaids who are working in registered kindergarten and nurseries in Jordan is crucial to providing and improving care.This study was the first of its kind in the region that investigated babysitters’ and nursemaids’ knowledge about Infant BLS and provided basic information which can be used to construct more appropriate and effective intervention programs.

### Aims

This study was designed to assess the level of awareness and the attitudes caregivers held towards the practice of BLS for an infant, among registered nurseries in north rural Jordan.

## METHODS

A cross-sectional study conducted during February 2017, child caretakers (babysitters and nursemaids) who were looking after infants in registered nurseries in north state of Jordan. A semi-structured questionnaire was distributed among babysitters and nursemaids who agreed to participate in the study. The questionnaire explored their knowledge, attitudes and perceptions regarding Infant BLS.

### Instruments

A semi-structured questionnaire was used, which was adapted from a study conducted by Chia and Lian (2014) in Singapore to assess parental knowledge, attitudes, and perceptions towards IBLS.(15) The questionnaire is to collect information about the demographic profile of the participants; knowledge for participants on IBLS skill, and questions of perceptions and attitudes about infant BLS. The questionnaire contains multiple options, True or False questions, and open ended questions. (15)

### Ethical Considerations

This study was approved by the concerned institutional review board. All participants signed a consent form after had information about the study and they have been notified that their participation was voluntary and they could withdraw at any time. They were guaranteed anonymity and confidentiality

### Data Analysis

Data were analyzed using SPSS 19 (SPSS, Inc., Chicago IL, USA). Descriptive statistical analysis was used to identify frequencies and percentages of studied categories, and corrolelation between demographical proflile and knowledge regardibg BLS.

## RESULTS

A total of 77 babysitters and nursemaids from an eighteen registered nurseries were recruited to participate in the study. The demographic characteristics are presented in Table-1. As seen in the table, approximately half of the participants have a diploma degree (52%), while another third (34%) has a bachelor’s degree, with only 14% having a secondary or less degree of education. In terms of age groups, around half of the participants (47%) were between 26 to 35 years old, while a quarter (26%) were between 36 to 45 years old, followed by (21%) of 25 years or less, and only (6%) were more than 45 years old. Lastly, concerning nationality, the majority (70%) were of Jordanian nationality, while the rest (30%) were refugees or migrants.

**Table 1:** The demographic profile of the participants (N=77)

| Characteristic | Sub-factors | n | % |
| --- | --- | --- | --- |
| education | Do not hold college/university degree | 28 | 36% |
|  | Hold college/university degree | 49 | 64% |
| Age group | 20-35 years | 52 | 68% |
|  | 36-50 | 25 | 32% |
| Nationality | Residents | 54 | 70% |
|  | Not resedent (Refugees/Migrant) | 23 | 30% |

### Babysitters’ and Nursemaids’ Knowledge of IBLS

The study investigated babysitters’ and nursemaids’ knowledge of the guidelines for IBLS, according to the American Academy of Pediatrics and the American Heart Association; including foreign body aspiration and CPR. Table-2 presents the responses related to participants’ knowledge regarding IBLS and foreign body aspiration. Based on babysitters’ and nursemaids’ responses, more than a third of them (38%, n= 29) never heard about CPR for children. Furthermore, the vast majority of participants (70%, n= 54) did not attend the training/education session regarding managing foreign body aspiration. However, 84 percent (n= 65) indicated that they would welcome participating in IBLS if it is offered to them.

**Table 2:**
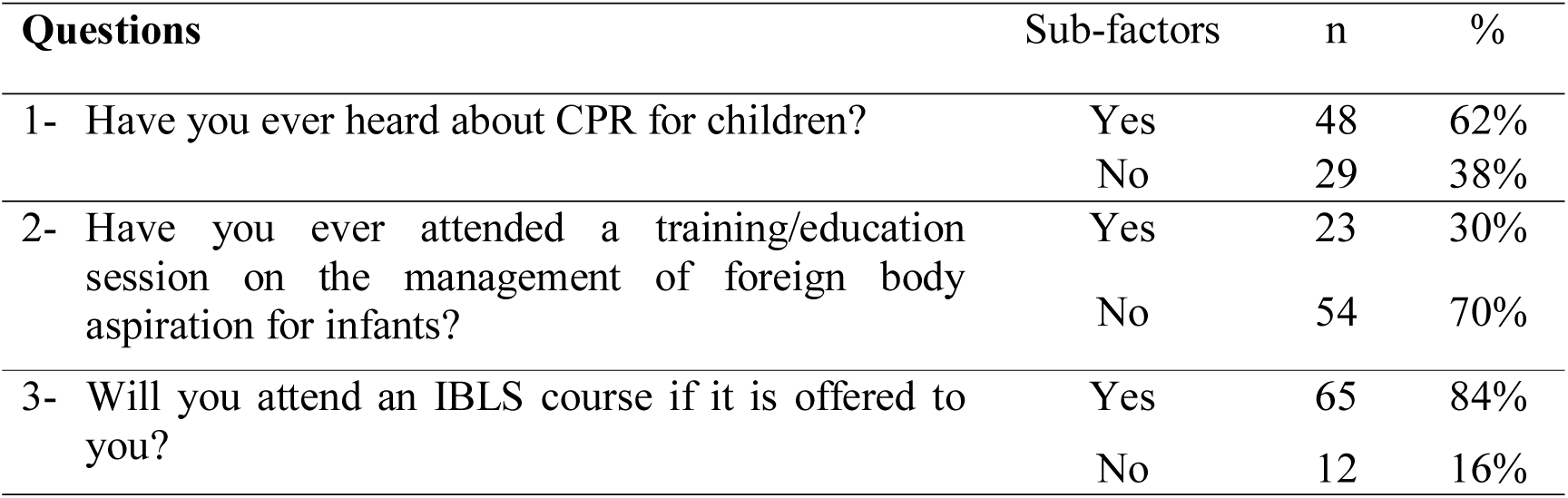
Knowledge regarding Infant basic life support/ foreign body aspiration

### The Proportion of Participants Who Obtained Passing Marks on General Information for BLS

As shown in Table-3, more than one-quarter (27%) of the participants were unaware of the Jordanian ambulance access number. Furthermore, only (66%) of nursemaids and babysitters answered correctly in relation to the child’s brain survival without oxygenation (breathing). When asked about the location to check a pulse in a child aged less than one year old, only 22% could identify the correct answer.

**Table 3:** The proportion of participants who obtained pass marks

| Question | Correct answer | % of correct responses |
| --- | --- | --- |
| 1. What is the Jordanian Ambulance access number? | 199 | 73% |
| 2. How long can a child's brain live without oxygen (breathing)? | 4-6 minutes | 67% |
| 3. Where is the location to check a pulse in a child aged less than 1 year old? | On the brachial artery (inside of the upper arm between the elbow and the shoulder) | 22% |
| 4. I can try to wake an unresponsive infant by: | Gently tap or flick the infant's feet soles | 44% |
| 5. I can confirm that an unconscious child is not breathing by: |  |  |
|  | Looking for rising and fall of the chest. | 48% |
|  | Listening for air escaping during exhalation. | 29% |
|  | Feeling for the flow of air from the infant's mouth and nose using my cheek. | 12% |
| 6. the correct ratio compression to breath to provide effective CPR for an infant |  |  |
|  | 30 compressions: 2 breaths if you are alone as rescuer | 13% |
|  | 15 compressions: 2 breaths if there are two rescuers | 39% |
| 7. If you witness a conscious infant unable to breathe and choking from a foreign body aspiration, what you should do to save his life? | Provide infant of 5 back blows and 5 chest thrusts until the object is expelled | 21% |
|  | Remove the piece aspirated by my fingers, only if I can see it | 43% |

Additionally, when asked the nursemaids and babysitters about the methods they may use to wake an unresponsive infant, only 44% answered correctly and indicated that they need to tap or flick the infant’s feet soles gently. Finally, participants were asked about how they can confirm if the baby is not breathing, about 48% of the participants answered the first item correctly, while correct answers composed 29% for the second item, and 12% for the third item (see Table-3).

With regards to their knowledge of the correct ratio of chest compression to breath rescue during BLS, only 13% of the participants were aware of the correct ratio compression to breath to provide an effective CPR for an infant with only one rescuer, and 39% indicated the correct answer for providing effective CPR for an infant when two rescuers are available (see Table-3).

As shown in Table-3, the questionnaire further investigated participants’ knowledge about proper interventions to be done in case of infant foreign body aspiration. Four alternative answers were provided with the right answer being “Provide infant of five back blows and five chest thrusts until the object is expelled,” only 21% participants were able to provide correct answer. Another 43% indicated that they should remove the piece aspirated if only were able to see it.

### The Main Source for the Information about IBLS

The majority of study participants, (52%, n=40), indicated that social media was their main source for information regarding BLS. While less than a quarter (21%, n=16) indicated that they learned about IBLS from conferences and training workshops, and only (16%, n=12) learned about it from friends and colleagues. Of the participants, few (12%, n= 9) reported that they access professional journals.

### Perception for Confidence Level among Babysitters and Nursemaids Regarding IBLS Skills

The study sample varied in their responses regarding their confidence in providing IBLS and dealing with foreign body aspiration situations. A high proportion of babysitters and nursemaids reported being not confident (42%) or not confident at all (22%). While only 30% of babysitters and nursemaids reported being somewhat confident, and just 6% were very well confident of being able to provide IBLS.

Contrarily, when they were asked about their ability to manage foreign body aspiration in infants, 35% of participants reported being not confident, and another 13% reported being not confident at all. Those who were somewhat confident composed 42% of the babysitters and nursemaids, while only 10% were very confident that they could provide proper intervention if such emergent event happened (see Table-4).

**Table 4:** Perception about confidence in providing IBLS/ foreign body aspiration management

| Questions | Count (%) |  |  |  |  |
| --- | --- | --- | --- | --- | --- |
|  | Not confident at all | Not confident | Somewhat confident | Very well confident | Total |
| Do you feel confident to provide a CPR for an infant in such an urgent situation? | 17 (22%) | 32 (42%) | 23 (30%) | 5 (6%) | 77 (100%) |
| Do you feel confident about your current knowledge of managing foreign body aspiration among infants? | 10 (13%) | 27 (35%) | 32 (42%) | 8 (10%) | 77 (100%) |

Furthermore the level of awareness among the babysitters/nursemaid regarding infant BLS was significantly (p<0.01) associated with level of education and age. Babysitters/nursemaid with no university/instition degrees were have less knowledge than babysitters/nursemaid holding college/university degrees. As well babysitters/nursemaid of ageing more than age of 35 have better knowledge, while there is no relation with residencey status (p>0.01). (Table-5)

**Table-5:** Correlations.

|  | knowledge |
| --- | --- |
|  | Pearson Correlation Sig. (2-tailed) |
| **Education** | .468(**) |
|  | .000 |
| **Age** | .451 (**) |
|  | .000 |
| **Residency** | .092 |
|  | .734 |
| N | 77 |

\*\* Correlation is significant at the 0.01 level (2-tailed).

### Challenges for Attending IBLS Courses

The study participants were asked to indicate their most significant challenge for attending such BLS training courses. The study participants indicated that the major challenge they face for attending IBLS courses was a time limitation due to overload work and busy schedule at nurseries. They also indicated that the long day shift prevent them from engaging in such course, some participants indicated that they only have a one day off work per week and so it difficult to attend these training events. In addition, participants indicated financial difficulties, and transportation difficulties were a challenging factor for attending the IBLS course.

### Suggestion for getting the training

The participants were also asked to choose their more significant way they prefer to receive such training or information about BLS. Less than quarter (21%) preferred the online training courses if it offered. Another 33% chosen face-to-face training, while 53% indicated that they prefer both ways to get the knowledge about BLS, another 3% were unsure about the method to receive such training.

## DISCUSSION

This study was conducted among 77 babysitters and nursemaids with the majority having a bachelor’s or diploma degree. The majority of babysitters and nursemaids were not aware of skills for managing foreign body aspiration among infants, and more than two-thirds of them never attended training or education sessions regarding managing foreign body aspiration and CPR practicing among children. However, the majority indicated that they are motivated to participate in such an event if it is offered to them. Significant differences were found in a previous randomized controlled trial study conducted in a tertiary pediatrics’ hospital in UK, which aimed to compare the infant CPR technical skill performance among several groups— one which participated in a self-motivated CPR refresher course using a competitive leaderboard, another with a CPR training feedback device, and a final group who did not participate in the course. The results showed that those who chose to participate in the refresher course by self-motivation scored significantly higher than the other groups. The study concluded that self-motivated participation in infant CPR training would help to improve knowledge and skills to manage infants in a critical situation.(16) Furthermore, the level of awareness among the babysitters/nursemaid regarding infant BLS was associated with the level of education and age, while there is no relation with residency status. Babysitters/nursemaid with college or university education level could have a chance of getting information about life support skills through meetings or studying. Also, babysitters/nursemaid with higher ages may have more life experiences.

One of the alarming findings of the current Jordanian study is that almost two-thirds of the proportion of babysitters and nursemaids were not confident in performing CPR for the infants in case of emergency. Furthermore, they lacked knowledge about the guidelines that they should follow in managing foreign body aspiration among infants. Babysitters and nursemaids were also unaware of how to confirm that a child is not breathing when is unconscious. There is insufficient training or qualification for kindergarten staff, especially in rural areas, where factors such as distance from the city centre may play a role in reducing the opportunities for continuous training of nursery staff. The economic factor could be a challenge, transport costs and training fees may influence the opportunity to get babysitters involoved in such training courses.

A descriptive study was conducted by Chia and Lain (2014), utilizing a survey questionnaire for knowledge, attitudes, and perceptions of parents in Singapore about IBLS.(15) The results of the study showed that IBLS training, as part of basic cardiac life support training, is important given that CPR can significantly alter the outcome in children with cardiopulmonary arrest. The study revealed that knowledge gaps can be filled through official training. It recommended a refreshing course regularly update ’ knowledge and skills among child caregivers.(15) Developing such training intervention for babysitters and nursemaids is highly needed in Jordan. Education campaigns have succeeded in part, and primary health care providers can play a crucial role in increasing education efforts about the risk of suffocation and management measures.(17)

Collaboration between the ministry of health, the ministry of education, the ministry of social development and civil defense in creating programs and policies for enhancing awareness of basic life support skills would be beneficial in accomplishing the mission. More than half of babysitters and nursemaids in this study indicated they use social media to gain information regarding basic life support skills. Using the Internet and adapting online training courses to address this need would be beneficial for targeting babysitters and nursemaids providing a suitable time for them to access the information, as well as being cost-effective by eliminating transportation needs. Value is added when thinking about further benefits of online training courses, such as increasing candidate independence, lowering teacher burden, and maintaining better standardization of course materials; additionally, it would be effective in terms cost.(18) However, first aid online courses may be useful for acquiring knowledge, but maybe not effective for performing BLS skills.(19) Nonetheless, pre-training evaluation of care providers knowledge and skills may be particularly beneficial in improvement BLS skills acquisition immediately after the course of the training.(20) Officials, governmental decision-makers, and stakeholders should support such initiative programs

The present study had a limitation with regard to the small sample size which affects general claims about all of the babysitters and nursemaids in Jordan. However, conducting this research was highly needed in Jordan, as far it was the first of its kind in the region that investigated caregivers’ (babysitters and nursemaids) knowledge about infant BLS, and provided basic information that used in constructing such an intervention program.

## CONCLUSION AND RECOMMENDATIONS

A majority of the study participants had not attended any CPR or BLS courses. However, it was encouraging that most of them expressed an interest in attending such a training session in the future. It is highly needed that administers run continuous education and training programs on IBLS for babysitters and nursemaids and to provide them with updated information and keep them aware of best practices for dealing with an urgent situation, such as choking that may often occur among young children. Policy changes are needed so that each kindergarten will include a child nurse at each setting; so nurses can lead the development of regular in-service training to improve caregivers’ skills through their participation in IBLS courses. This regular and continuous scheduling will minimize any challenges that prevent caregivers form attending IBLS course including time limitation, transportation issues, and financial difficulties. Further, providing such Internet-based resources would help eliminate barriers like cost and transportation. Officials, governmental decision-makers, and stakeholders should support such initiative programs.

A special effort could be made to engage the newly graduated students by including them in such repairing courses before they begin working in the nurseries and kindergartens. At the community level, local community basic service centers, non-governmental organizations, local associations, universities and health colleges could support awareness through providing courses that aimed at enhancing knowledge and skills for basic life support among child caregivers and babysitters

## Data Availability

data available if the reviewer requested

## Acknowledgment

Acknowledge for AABU and JUST universities for supporting this project. Thanks to caregivers who accepted to take part to participate in the study survey.

